# Infectious, Allergic, and Immune-Mediated Disease Data Resources: A Landscape Overview and Subset Assessment

**DOI:** 10.1101/2025.07.30.25332458

**Authors:** Darya Pokutnaya, Lisa M. Mayer, Sydney Foote, Meghan Hartwick, Sepideh Mazrouee, Willem G. Van Panhuis, Reed Shabman

## Abstract

**Background:** The Data Management and Sharing (DMS) Policy issued by the National Institutes of Health (NIH) requires most grant applications to include a DMS Plan, detailing data type(s), resources (e.g., data repositories, knowledgebases, portals) for data sharing, and a dissemination timeline. Researchers face challenges navigating the complex data landscape to identify data resources to fulfill the DMS Policy requirements. The National Institute of Allergy and Infectious Diseases (NIAID) aims to support researchers in preparing DMS Plans for applications that align with its mission areas.

**Methods:** To support depositing and accessing infectious, allergic, and immune-mediated disease (IID) data, we compiled a list of IID data resources. The list was developed by reviewing online resources and collecting recommendations from subject matter experts. Additionally, we developed a questionnaire based on NIH recommendations and community best practices to characterize a subset of IID data resources that support data submissions.

**Results:** We identified 303 data resources, 58 of which focused on IID data. Most were categorized as General Infectious Diseases and Pathogens (n = 29), followed by Respiratory Pathogens (n = 10). Scientific content included “omics” (n = 37), clinical (n = 21), and biological assay data (n = 20). Open access data was common (n = 39), with fewer offering controlled access (n = 20) or required registration (n = 4). Among 19 resources accepting data submissions, eight required registration, seven needed additional approvals, and four required network membership. Fifteen resources provided metadata access, with 11 assigning persistent identifiers. Twelve offered APIs, 13 provided analytical tools, and 10 featured workspaces. Risk management documentation was available for 10, and five provided data retention policies.

**Conclusions:** We assessed 58 data resources in the IID domain, identifying 19 that support data submission and are therefore suitable for NIH DMS Plans. Our findings reveal both the breadth of available resources, and the challenges related to inconsistent data submission requirements and data management practices. Enhancing transparency and standardization across data resources will support more effective data sharing, enhance findability, and aid researchers in selecting appropriate resources for DMS Plans and secondary data analysis.

## Background

To promote the transparency, accessibility, and usability of scientific data, the National Institutes of Health (NIH) implemented the Data Management and Sharing (DMS) Policy (1). The policy requires a DMS Plan for most grant applications that describes the data resource(s) (e.g., data repositories, knowledgebases, portals) where data derived from the corresponding project will be deposited. Navigating the landscape of data resources, including generalist resources that accept a wide variety of data types and domain-specific resources that accept data from a particular field, is a time-consuming task (2).

The NIH provides several recommendations for choosing appropriate data resources to include in a DMS Plan, including considerations of scientific discipline, data type, volume, and long-term access, storage, security, and reuse (3–5). These characteristics align with frameworks such as the FAIR (Findable, Accessible, Interoperable, Reusable) and TRUST (Transparency, Responsibility, User Community, Sustainability, and Technology) Principles, CoreTrustSeal requirements, Office of Science and Technology Policy (OSTP) Desirable Characteristics of Data Repositories for Federally Funded Research, as well as other domain-specific repository evaluations (3, 6–9). The NIH also offers several discipline-specific tools, including the National Cancer Institute Data Catalog and the National Institute of Child Health and Human Development Data Repository Finder. Despite these recommendations and tools, there is currently no dedicated tool for identifying data resources specific to infectious and immune-mediated disease (IID) research. As a result, IID researchers continue to face challenges in identifying data resources that align with the NIH guidelines outlined in the DMS Plan.

Given this lack of targeted tools for IID research, deciding where to submit data can be an obstacle for researchers. These decisions are often influenced by multiple factors, including data use limitations, the types of data being shared, sustainability of the resource, and the ease with which others can discover and use the data. Many biomedical researchers lack formal training in informatics, making it difficult to evaluate resources effectively. As such, tools are needed to support researchers in making informed choices about data sharing. The assessment and questionnaire presented in this manuscript can serve as a valuable example of how researchers might assess potential resources for hosting their data.

Our goal is to characterize data resources to assist IID researchers deciding where to deposit data. Recognizing the complexity of this decision, including factors such as data submission requirements, metadata standards, and access features, we aim to 1) describe data resources that store IID data, 2) provide resources to help researchers develop DMS Plans, and 3) highlight resources that contain datasets suitable for secondary analyses.

We identified 58 IID-specific data resources and conducted a comprehensive assessment of 19 that support data submission using a questionnaire developed in this study. Our assessment focused on key attributes that support long-term data access, storage, security, and data reuse, as these features enhance data accessibility, usability, and interoperability. This assessment offers an overview of the current IID data resource landscape, providing valuable insights to support the NIH and NIAID’s mission to maximize data sharing and reuse to advance the understanding, treatment, and prevention of infectious, allergic, and immune-mediated diseases.

## Methods

### Initial landscape review of infectious, allergic, and immune-mediated disease data resources

Data resources were identified between November 2022 and March 2025 through a curated, expert-informed review of publicly available websites and in consultation with NIAID-affiliated subject-matter experts (SMEs) including data scientists, Program Officials, and NIAID-funded researchers involved in the generation, management, or storage of IID data (Fig. 1). The resources included were not limited to NIH- or NIAID-funded resources.

**Figure 1.**
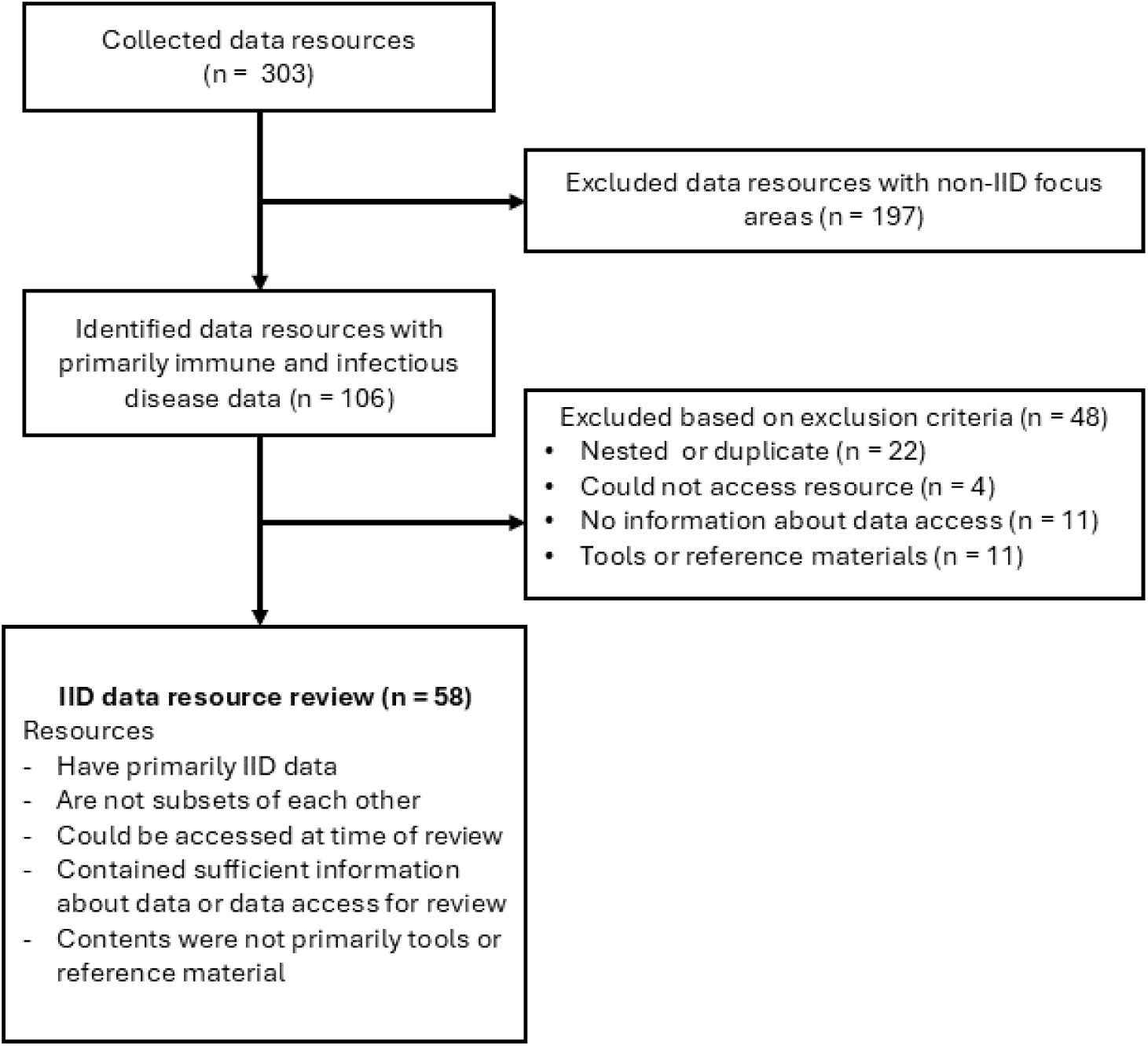
Infectious and immune-mediated (IID) data resource inclusion and exclusion criteria workflow.

Data resources classified as having primarily IID data were evaluated for exclusion criteria. Resources were excluded if they were nested within larger resources, if they were no longer accessible, lacked information about data access, or if the resource only contained reference materials. The remaining resources were described further, identifying features such as the primary diseases or pathogens captured, scientific content, data access requirements, and data submission capabilities. Scientific content was categorized using terms and definitions based on the National Library of Medicine Medical Subject Headings (Table 1). Data access requirements were classified as either open, registration required, or controlled access based on established definitions (10). Each data resource was identified as either accepting or not accepting data submissions. Resources that accepted data submissions were subset for the questionnaire assessment.

**Table 1.** Data resource scientific content terms, definitions, and NLM MeSH entries referenced when developing the definitions.

| Scientific Content Term | Definition | MeSH Entries |
| --- | --- | --- |
| **Biological assay data** | Consists of data that measure the effects of a biologically active substance using an intermediate in vivo or in vitro tissue or cell model under controlled conditions | Biological Assay |
| **Biospecimens** | Represents tissue samples retained from their initial research or medical purpose in a biorepository | Biological Specimen Banks |
| **Clinical data** | Comprises patient data collected through medical care such as medical records and results from diagnostic techniques and procedures | Medical Records |
| **Epidemiological data** | Includes data from or related to studies in epidemiology (e.g., data related to causes, incidence, and characteristics behaviors of disease outbreaks affecting human populations) | Epidemiology |
| **Imaging data** | Consists of image data, such as outputs from diagnostic imaging or microscopy | Diagnostic Imaging |
| **Laboratory chemicals** | Describes chemicals used or produced in laboratory research, such as reagents or drug formulations | Laboratory Chemicals |
| **Metadata catalog** | Contains metadata describing datasets which are stored elsewhere | Metadata |
| **Omics data** | Contains data from genomics, proteomics, metabolomics, multi-omics, and related disciplines | Genomics, Proteomics, Metabolomics, Multiomics |
| **Software** | Includes downloadable software or computational tools | Software |

*Abbreviations: NLM, National Library of Medicine; MeSH, Medical Subject Headings. Note:* All definitions for scientific content terms were sourced from the *MeSH* database.

### Questionnaire development and subset assessment

Following our external assessment, a 23-item questionnaire was developed based on key criteria from the FAIR and TRUST Principles, Office of Science and Technology Policy (OSTP) Desirable Characteristics of Data Repositories, CoreTrustSeal requirements, and several domain-specific repository evaluations (3, 6–9). We sought consensus across these sources to reflect commonly recognized elements of high-quality data resources. Furthermore, to promote consistency and objectivity, questions were designed to avoid subjective language and be answerable with a clear yes/no based on publicly available documentation. The questionnaire was developed to help researchers identify suitable resources as part of their DMS Plans and report on key characteristics that may influence resource selection.

The questionnaire items were categorized into four groups: 1) Data Access and Submission, 2) Identification, Provenance and Quality Assurance, 3) Data Retrieval and Analytical Tools, and 4) Documentation and Compliance. Co-authors LM and DP independently reviewed the IID data resources that accept submissions by completing the questionnaire using publicly available documentation on the data resources’ websites. This included content accessible without logging in, as well as information available to users who created a free account using an email address. No direct communication with the data resource staff occurred. Discrepancies between reviewers were resolved through discussion.

#### Data access and submission

For each resource, data access and submission were classified using three categories. “Submission Allowed with Registration/Account” included resources that required users to register for an account or sign up for the platform before submitting data. “Submission Allowed with Additional Approval or Contracts” defined resources that require additional steps beyond registration, such as contracts, agreements, or formal approval like those from an Institutional Review Board (IRB). “Submission Allowed with Membership” applied to data resources that require users to join a specific network or program prior to data submission. In addition to these classifications and previously described data access requirements, the questionnaire captured whether resources provided open metadata, supported authentication of data submitters, enforced formatting and size limitations for data submission, and whether fees were associated with data deposition.

#### Identification, provenance, and quality assurance

Resources were categorized as using persistent, local, or no identifiers to describe their data. Persistent identifiers were defined as stable, long-term references to digital objects, which can include Digital Object Identifiers (DOIs) or Internationalized Resource Identifiers/Uniform Resource Locators (IRIs/URLs) (11). Local identifiers were defined as those that are only guaranteed to be unique within the resource itself. The questionnaire also assessed whether each data resource tracks the provenance of metadata and data, and whether there is expert curation or quality assurance support once the data are submitted.

#### Data retrieval and analytical tools

The third section of the questionnaire assessed the presence of data retrieval and analytical tools. Resources were evaluated based on whether the data or metadata could be accessed through an Application Programming Interface (API) or downloaded onto the user’s machine. It also evaluated whether the site provided any analytical tools or a dedicated workspace for analysis. If a workspace was available, additional questions addressed associated costs with maintaining or analyzing data and whether researchers could use their own tools within the workspace.

#### Documentation and compliance

The final section of the questionnaire assessed whether the data resources provided documentation on risk management, data retention policies, and security measures to protect against unauthorized access or modification based on data sensitivity. It also evaluated whether the resource outlined its terms of data use.

This review identified and described IID data resources, supported researchers in selecting appropriate repositories for DMS Plans, and highlighted resources that may have data suitable for secondary analyses. The assessment of IID resources that allow for data submission focused on attributes that promote long-term data access, storage, security, and reuse to enhance overall data accessibility, usability, and interoperability.

## Results

### Initial landscape assessment of infectious, allergic, and immune-mediated disease data resources

We performed a landscape assessment to identify data resources considered IID-specific. The complete list of the 303 data resources and website URLs is provided in Supplementary Table 1. After identifying the IID-specific resources from the 303, exclusion criteria were applied. A total of 48 resources were removed, including 22 nested resources, four that were inaccessible due to broken links, 11 that lacked available data or access guidelines, and 11 that contained only reference materials.

We summarized the subset of 58 IID data resources, including the primary disease or pathogens captured, scientific content categories, data access categories, and data submission acceptance (Table 2). Additional information such as resource abbreviations and URLs are found in Supplementary Table 2. Most data resources were categorized by their primary disease or pathogen as General Infectious Diseases and Pathogens (n = 29), Respiratory Pathogens (n = 10), and HIV/AIDS (n = 8) (Fig. 2 and Supplementary Table 3). The ChemDB HIV, Opportunistic Infection and Tuberculosis Therapeutics Database was categorized as having both Respiratory Pathogen and HIV/AIDS data. Five resources were categorized under Immunological and Autoimmune Diseases, and three under Hemorrhagic Fever Viruses. Arthropod-borne Pathogens, Aspergillosis, Papillomaviruses, and Hepatitis C were each classified as the primary disease or pathogen for a single data resource.

**Figure 2.**
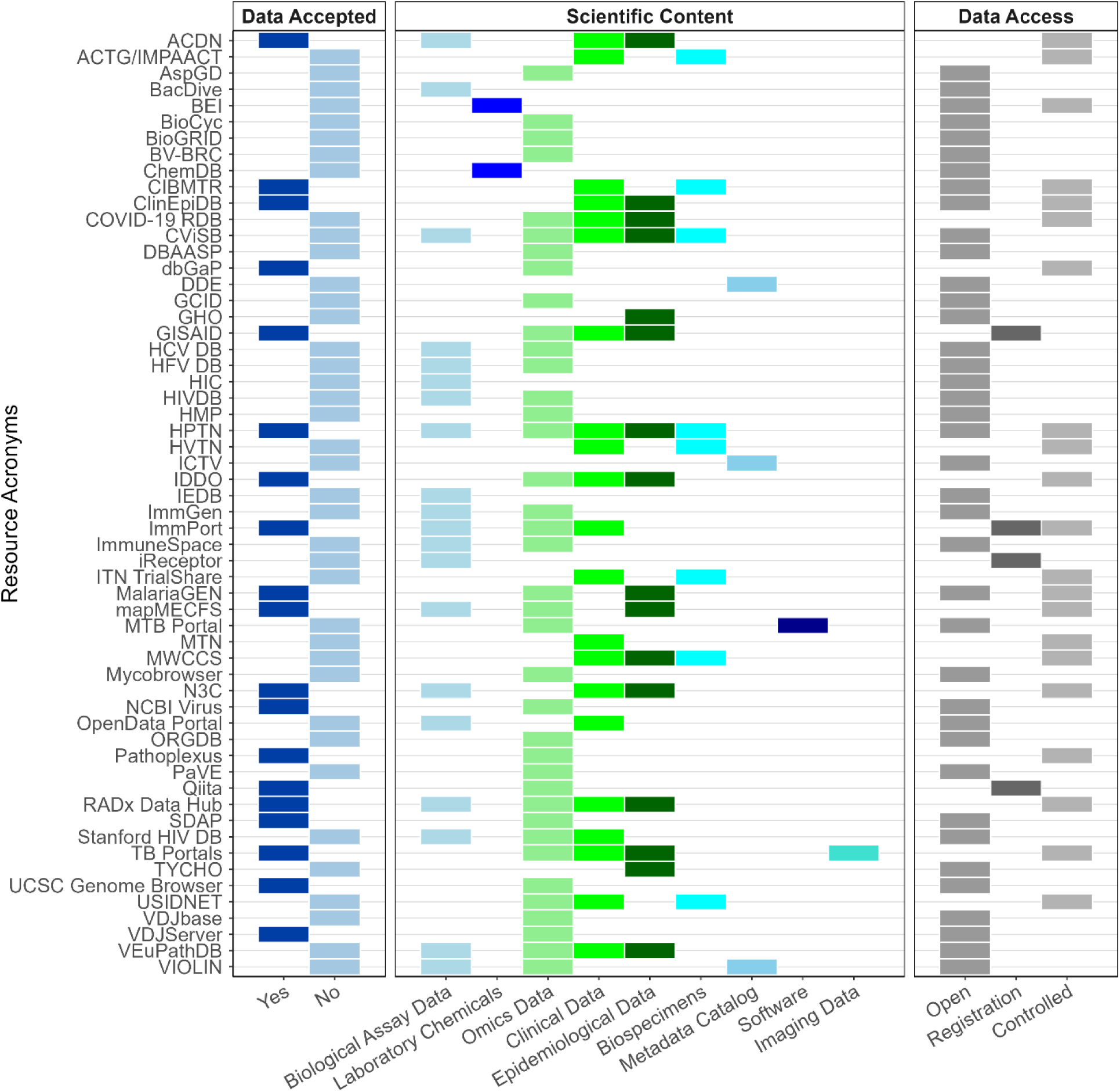
Data submission acceptance, scientific content categories, and data access categories of infectious and immune-mediated data resources (n = 58).

**Table 2.**
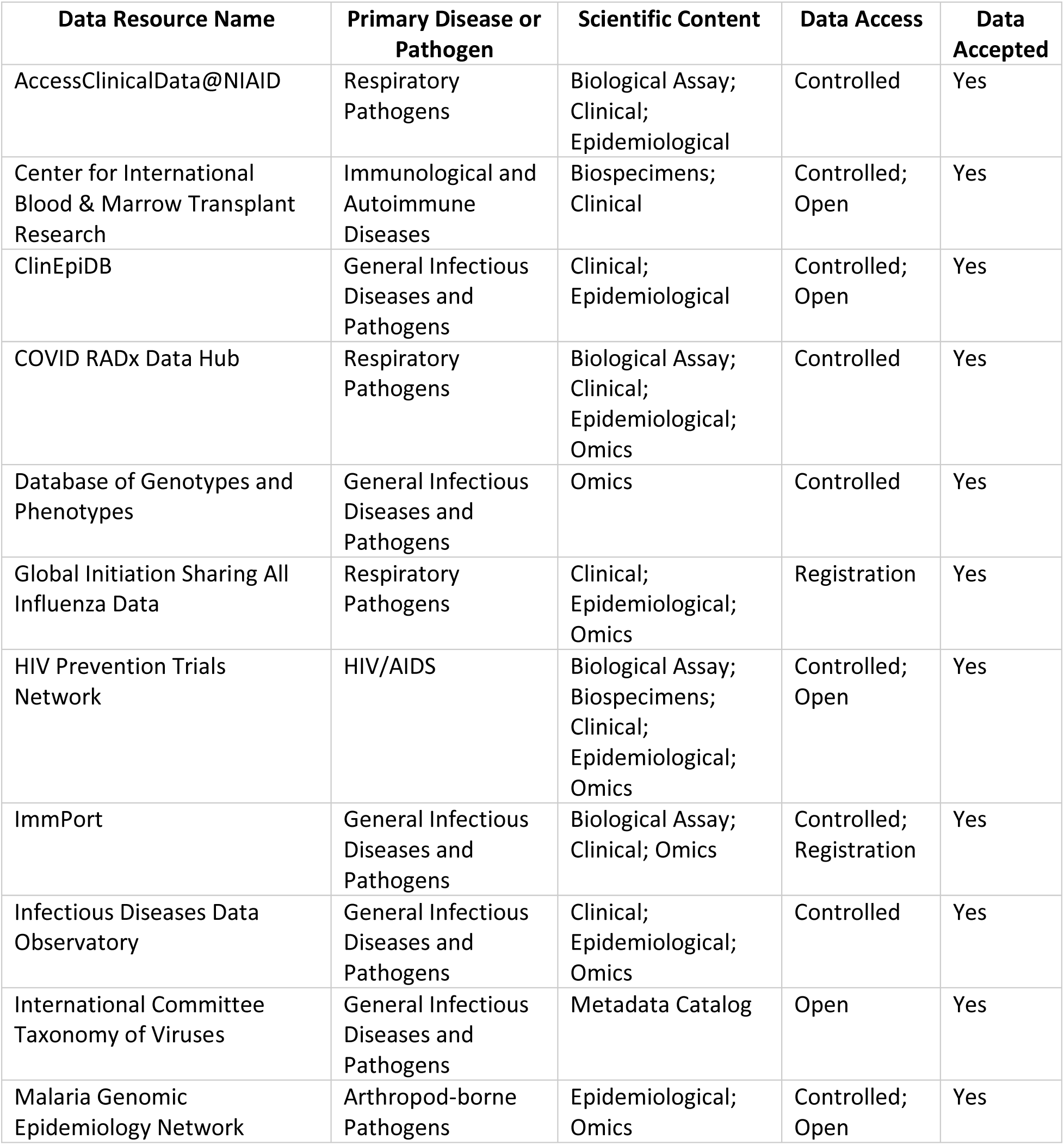
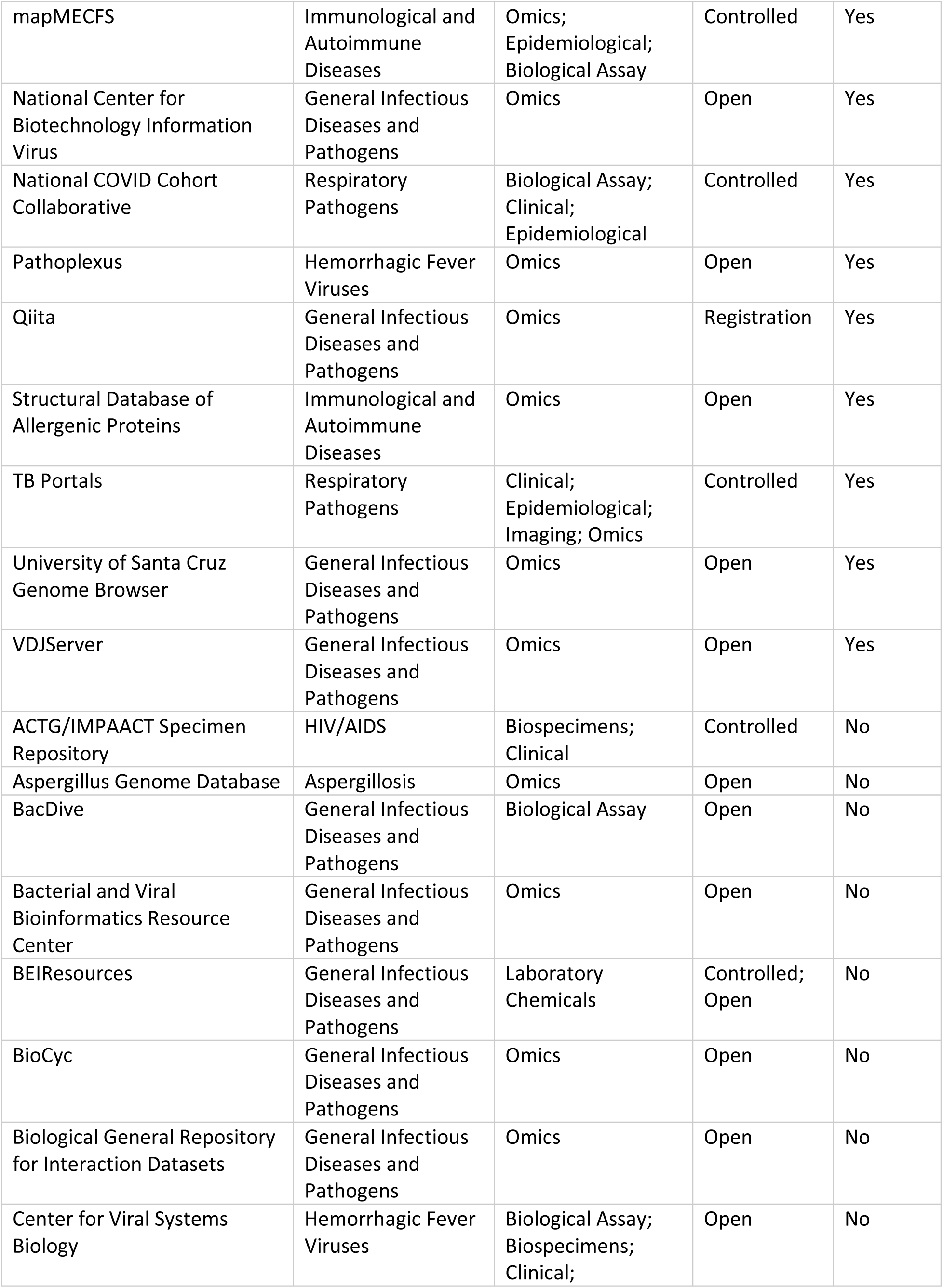
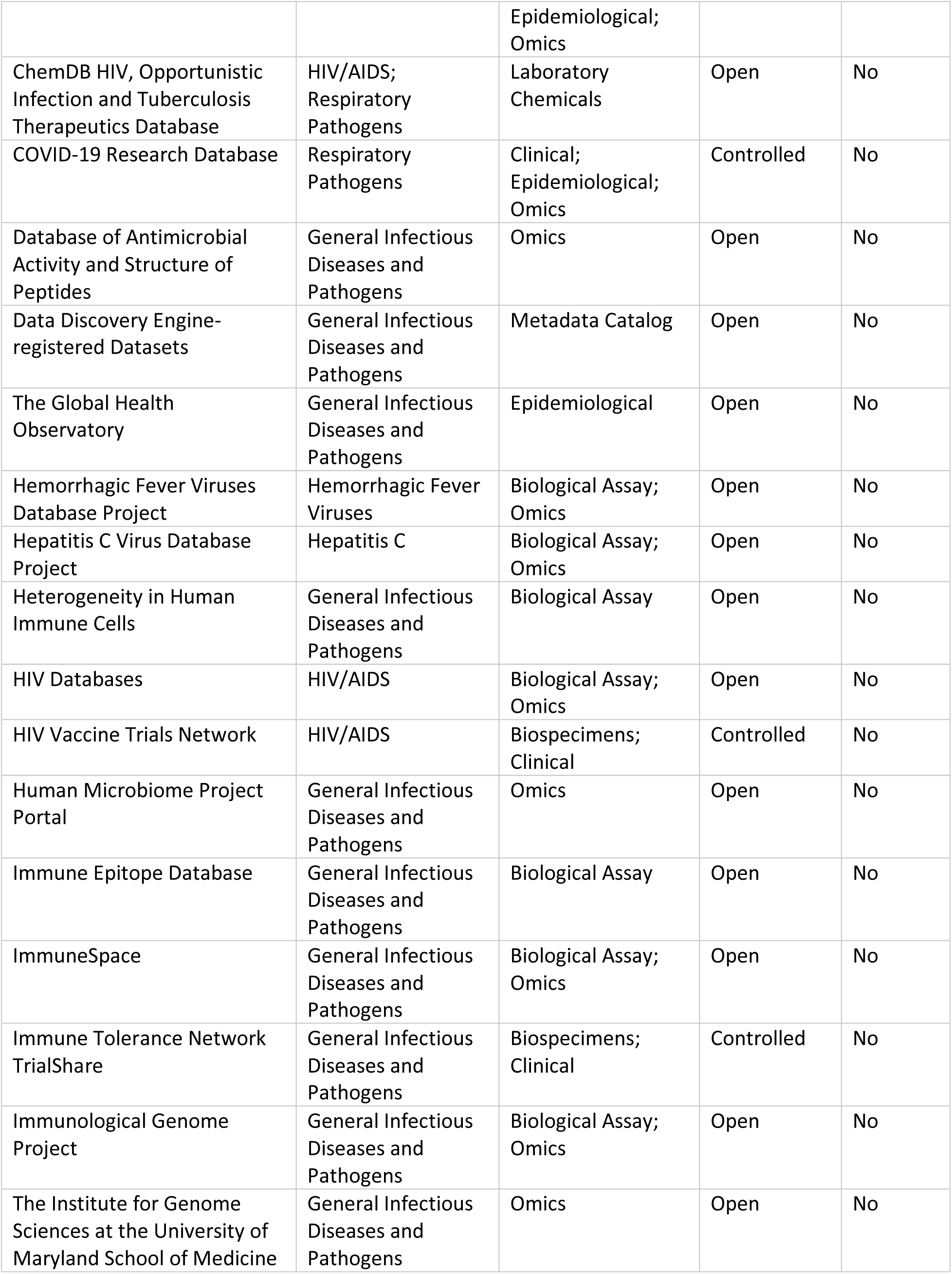
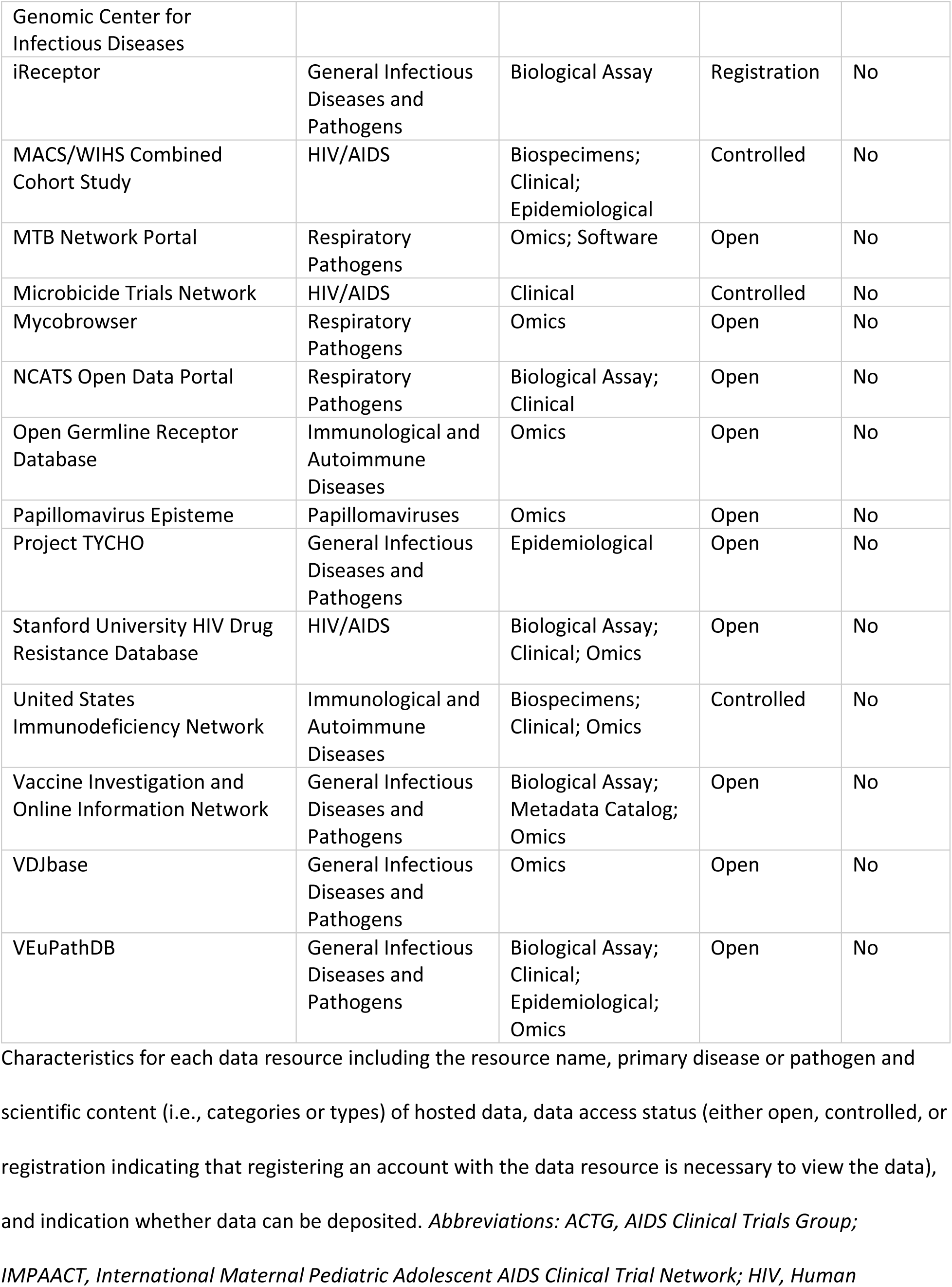
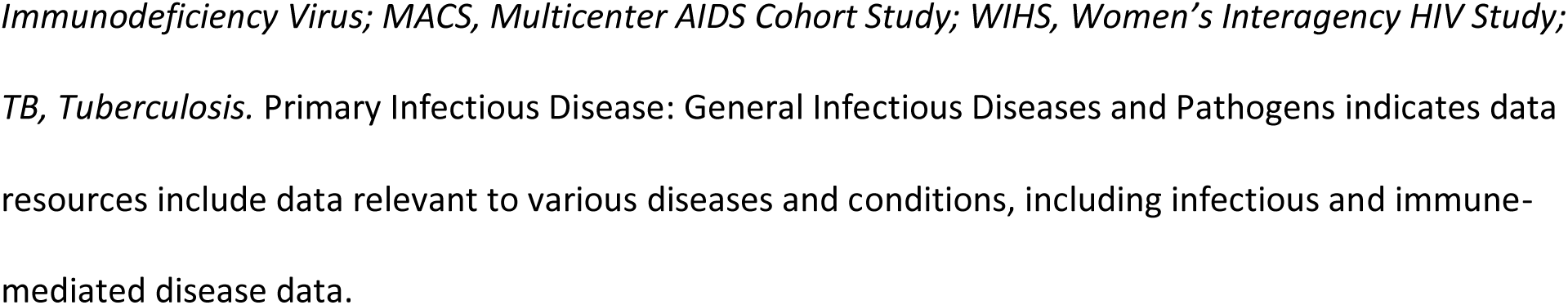
Core characteristics of identified infectious and immune-mediated (IID) data resources (n = 58) ordered by data submission acceptance and alphabetically.

### Resource classification by data access

Thirty-four of the 58 IID resources provided only open access data (Fig 2. and Supplementary Table 4). Fifteen resources contained controlled access data; access required additional measures beyond registration. Five resources offered both open and controlled access data. Three were classified as having registration only data, indicating that an account registration is required. One resource was categorized as having both registration and controlled access data, signifying that some data are available upon user registration, while others require additional steps for controlled access.

### Resource classification by scientific content and data submission acceptance

Data resources contained scientific content from one or more of the categories. Thirty-eight included “omics” data (e.g., genomics, proteomics, metabolomics, multi-omics, and related disciplines) with 15 allowing for data submission. Twenty-one contained clinical data (e.g., medical records and results from diagnostic techniques and procedures), of which ten accepted submissions. Biological assay data appeared in 20 resources, with six enabling user submission and 16 had epidemiological data, ten of which allowed for submission (Fig. 3 and Supplementary Table 5). Additionally, eight resources featured biospecimens, with two accepting data deposition. Three were categorized as metadata catalogs, two featured laboratory chemicals, one provided a list of software, and one contained imaging data, which was the only resource among these that supported data submission.

**Figure 3.**
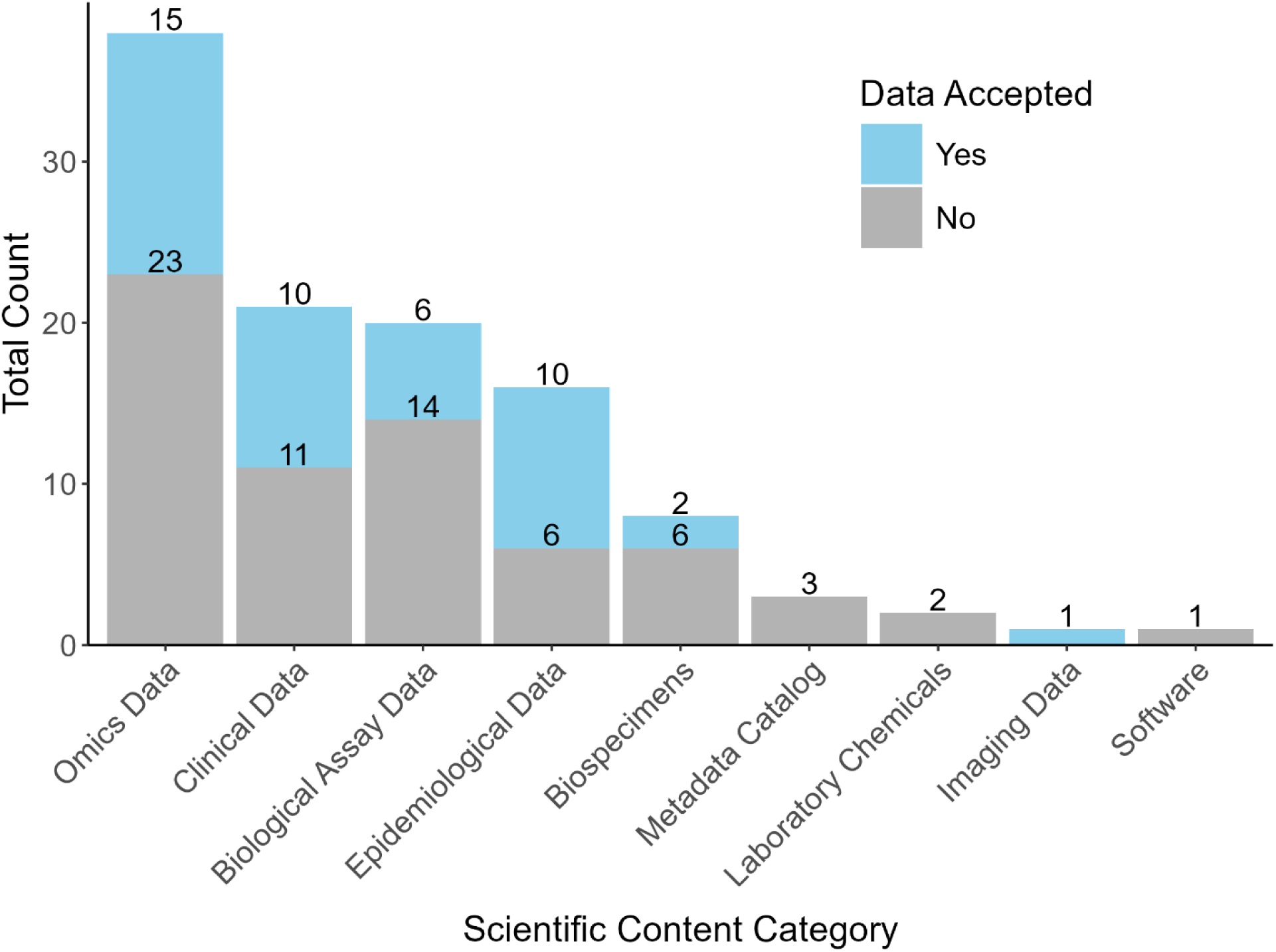
Counts of data resources (n = 58) by scientific content categories and data submission acceptance. Some resources are represented in multiple categories.

### Questionnaire and subset assessment

Out of the 58 IID data resources, we identified 19 that allow for data submission. Among them, eight required registration or an account prior to submission (Table 3). Seven required additional contracts or approvals, such as a data use agreement or IRB approval prior to accessing the data. Four required that researchers be part of a specific collaborative network or consortium to submit data.

**Table 3.** Aggregate evaluation results of infectious and immune-mediated disease data resources that accept data deposits.

| Category | # | Question | Response |  |  |
| --- | --- | --- | --- | --- | --- |
|  |  |  | Submission Allowed with Registration/ Account | Submission Allowed with Additional Approvals or Contracts | Submission Allowed with Membership |
| 1) Access and Submission | 1.1 | Does the resource accept data submission? | 8 | 7 | 4 |
|  |  |  | Yes | No | NA |
|  | 1.2 | Does the data resource provide open access data? | 8 | 11 | - |
|  | 1.3 | Does the data resource require registration (e.g., email) for data access? | 7 | 12 | - |
|  | 1.4 | Does the data resource provide controlled access data? | 13 | 6 | - |
|  | 1.5 | Does the data resource provide open access metadata? | 15 | 4 | - |
|  | 1.6 | Does the data resource support authentication of data submitters? | 19 | 0 | - |
|  | 1.7 | Does the data resource have formatting requirement for data submission? | 10 | 9 | - |
|  | 1.8 | Does the data resource have size limit requirements for data submission? | 1 | 18 | - |
|  | 1.9 | Are there costs associated with depositing the data? | 1 | 18 | - |
| 2) Identification, Provenance, and Quality Assurance |  |  | Persistent Identifier | Local Identifier | No Identifier |
|  | 2.1 | Does the data resource assign each dataset an identifier? If yes, is it a persistent or local identifier? | 5 | 11 | 3 |
|  |  |  | Yes | No | NA |
|  | 2.2 | Does the data resource have a system in place to track provenance to the (meta)data? | 14 | 5 | - |
|  | 2.3 | Does the data resource support expert curation or quality assurance to improve the accuracy and integrity of datasets and metadata? | 14 | 5 | - |
| **3) Data Retrieval and Tools** | 3.1 | Can the (meta)data be accessed through an API? | 12 | 7 | - |
|  | 3.2 | Can the user download the data to their local machine? | 19 | 0 | - |
|  | 3.3 | Does the data resource provide data analytical tools? | 14 | 5 |  |
|  | 3.4 | Does the data resource provide a workspace? | 10 | 9 |  |
|  | 3.5 | If there is a workspace, are these costs associated with maintaining data in the workspace? | 1 | 9 | 9 |
|  | 3.6 | If there is a workspace, are users able to utilize their own analytical tools within the workspace? | 2 | 8 | 9 |
|  | 3.7 | If there is a workspace, are there costs associated with analyzing the data in the workspace? | 0 | 10 | 9 |
| **4) Documentation and Compliance** | 4.1 | Does the data resource provide documentation on risk management (e.g., data breach, natural disasters)? | 10 | 9 | - |
|  | 4.2 | Does the data resource provide documentation on its data retention policies? | 5 | 14 | - |
|  | 4.3 | Does the data resource have security policies in place that ensure protection against unauthorized access, modification, or release of data, with appropriate security levels based on data sensitivity? | 17 | 2 | - |
|  | 4.4 | Does the data resource provide documentation for its terms for data use? | 18 | 1 | - |

### Data access and submission

Seven of the 19 data resources accept data submissions with registration or an account, seven accept data submission with additional approval or contracts, and five require membership before submission. Six resources provide open access data, six require users to register, and 12 offer controlled access data. Regardless of data access, most data resources (n = 14) provide access to at least some metadata. All data resources support authentication of the data submitter. Nine of the resources have public-facing documentation on formatting requirements for data submissions. One data resource specifies a size limit requirement, and one charges a fee for data deposition.

### Identification, provenance, and quality assurance

Five data resources assign persistent identifiers (e.g., DOIs) to each dataset, 11 resources use local identifiers specific to their platform, and three resources do not assign any dataset identifier. Most data resources (n = 14) have a system in place to track provenance of the metadata or data. Most resources (n = 14) also support expert curation and quality assurance to improve the accuracy and integrity of the data and metadata.

### Data retrieval and analytical tools

Out of the 19 data resources evaluated, 12 provide access to the metadata or data through an API. All resources allow users to download data to their local machine. Thirteen offer at least one analytical tool, while nine provide a workspace. Among the data resources with workspaces, one requires payment to maintain data and two allow users to utilize their own analytical tools within the workspace. Notably, none charge a fee to analyze data in the workspace.

### Documentation and compliance

Nine data resources provide documentation on risks such as data breaches and natural disasters. Documentation on data retention policies is provided by five resources. Security policies ensuring protection against unauthorized access, modification, and release of data, with appropriate security levels based on data sensitivity, are in place for 16 data resources. All resources provide some documentation outlining their terms for data use.

## Discussion

Our assessment highlights the diversity and complexity of IID research, reflected in the wide range of data resources available. Selecting appropriate data resources for a DMS Plan is challenging and requires consideration of data types and formats, security, storage, retention policies, and the trade-offs between using a single or multiple resources for data deposition (2). These decisions affect data reuse, particularly if the resource is not widely recognized or lacks interoperability features (12). Early resource selection in the study process influences data and metadata formatting, access, and sharing policies. For example, repository requirements may influence informed consent language (13,14). These considerations underscore the need to integrate data resource selection not only during DMS Plan preparation, but even earlier during the study design phase so that data management strategies are aligned from the outset and can support future sharing and reuse effectively.

Given the importance of selecting appropriate data resources early in the study design, our assessment highlights an imbalance in the availability of resources for IID research. While “omics” and clinical data are well-represented, other categories including imaging and biospecimens are notably underrepresented. These imbalances may stem from differences in investment, data sharing culture, and technical or ethical challenges. For example, the bioinformatics community was among the first to embrace data sharing, leading to the development of specialized “omics” resources (9,15). In contrast, imaging data requires significant storage space and is difficult to de-identify (16). Sharing biospecimen data presents unique challenges due to need for strict ethical oversight, governance structures, and compliance with clinical and laboratory standards (17). As a result of these complexities, fields outside of “omics” may have fewer specialized resources available to support data deposition and long-term access. Although generalist resources such as Zenodo and FigShare remain options, domain-specific resources are better suited to support IID researchers by organizing data in ways that maximize discovery and utility.

We observed considerable variation in submission processes, access controls, metadata practices, and documentation quality in our subset assessment of 19 IID data resources that support data deposition. Data access models ranged from open to controlled, and authentication requirements varied from email registration to institutional approval. These variations in access and authentication have implications for both data reuse and DMS Plan development. While important to ensure data is appropriately protected, controlled access may delay reuse and secondary analyses. Complex authentication for or institutional restrictions on data deposition can also complicate resource selection and should be considered early by researchers to ensure compliance and feasibility (18).

Despite variation in data access, 15 of the 19 resources (Table 3) provided open-access metadata, enabling researchers to assess data relevance, structure, and quality before initiating access requests. This transparency is especially valuable for planning secondary analyses and selecting suitable resources during proposal development.

Data submission requirements varied across resources. In some cases, resources did not publicly provide guidance on file formatting and size. Researchers are asked in the DMS Plan to specify which standards, if any, will be applied to the scientific data and associated metadata, including data formats, data dictionaries, unique identifiers, and other documentation (1). However, this can be difficult to address when a data resource does not provide clear guidance on formatting, as the researcher is trying to align the data and metadata required for their study with resource guidelines.

Providing clear details on submission requirements may save researchers time by preventing them from having to retroactively adjust formatting if hidden requirements are discovered later in the process.

Practices for assigning dataset identifiers varied across resources. While some resources issued globally unique identifiers such as DOIs, others used local identifiers that may not be resolvable outside their original context or interoperable across platforms. In contrast, DOIs support consistent data citation, long-term accessibility, and integration across resources. Aligning with the FAIR Principles and recent NIH and OSTP guidance, resources are increasingly expected to assign unique, citable persistent identifiers to support access and tracking of federally funded research (3,9,19).

These differences highlight the importance of reviewing resource documentation before submitting a DMS Plan, and when needed, engaging directly with resource staff to ensure alignment with data sharing goals (20).

Provenance, or the origin and history of the data, differed by resource. We considered a resource to support provenance tracking if it publicly documented any aspect of data versioning. This ranged from systems where users manually updated files with version numbers and deposit dates to those with automated provenance tracking. Automated systems are significantly more reliable and consistent than manual methods, which are prone to human error (7). While variation in identifiers, tracking systems, and submission requirements may present challenges, they also offer researchers flexibility in selecting resources that best align with their data types, access needs, and management goals.

The variability in features highlights the importance of active support within resources to help researchers manage their data efficiently and prepare for DMS Plan submission. Fourteen of the 19 resources provided expert curation or quality assurance practices that support improvements to data and metadata post-submission (Table 3) (21). These practices not only support compliance with data sharing policies but also promote greater confidence in the reliability and reusability of shared data.

All resources allowed users to download data locally, and a majority supported access through APIs, offering flexibility in how metadata and data are accessed and integrated into workflows. Fourteen resources provided at least one built-in analytical tool, while over half offered workspace environments. Only one resource reported costs associated with maintaining data in the workspace, and none required payment for data analysis. These finding suggest that while workspace availability is not universal, they are generally low-cost and accessible when offered. However, only two of the resources with workspaces allowed researchers to utilize their own analytical tools. This limited flexibility in tool integration may influence resource selection for DMS Plans based on project-specific needs.

Documentation on risk management, data retention, and security policies was often difficult to locate and interpret across the 19 resources. While some level of risk management documentation, covering potential threats such as data breaches and natural disasters, was available, nine resources did not offer any. This disparity suggest that researchers may need to conduct additional assessments of risk management or reach out to data resource staff directly to ensure adequate protection against unforeseen events that could impact their data. Only five resources documented data retention policies, while the remaining 14 providing no clear guidance. This gap is important, as understanding data retention terms supports long-term project planning and data accessibility. Furthermore, DMS Plans require researchers to provide a timeline specifying how long scientific data will be available to others (1). In contrast, most resources provided some documentation on policies designed to protect data from unauthorized access. However, the phrase “appropriate security levels” in our assessment was interpreted broadly; we assumed that each resource’s verification process met the necessary security requirements unless documentation indicated otherwise. Most resources also included terms for data use, helping ensure that legal and ethical considerations for data sharing are clearly addressed.

### Limitations

Limitations in this assessment arise from reliance on public documentation, flexible handling of variation between resources, and changes in resources over the course of the evaluation. We relied solely on publicly available documentation, which may not capture all information about each resource (22). This limitation is inherent in the process that researchers also face when selecting a data resource for their DMS Plan or secondary analysis. Additionally, due to inconsistencies in documentation, the reviewers took a flexible approach, giving credit to the data resource if any publicly available documentation was found for each question. Future work could assess each question along a spectrum rather than a binary response, to better understand the nuances in documentation and implementation of each resource. Finally, changes in funding or infrastructure may result in inactive links or outdated resources provided in the tables. For example, during the assessment, we evaluated the original version of VDJ Server, which was later deprecated and replaced by VDJ Server 2. We intend to create a dynamic, publicly available list of IID resources to ensure continued access to the latest information as changes occur.

## Conclusions

Our assessment differs from prior studies in two ways: 1) it focuses specifically on IID data resources, and 2) it assesses each resource by describing the presence of relevant features for DMS Plan development and secondary data analysis. The findings highlight the diversity and flexibility of resources available to researchers, spanning “omics,” clinical, epidemiological, and biological assay data, but also underscoring the significant challenges posed by variability in submission requirements and data management practices. These challenges emphasize the need for greater transparency and standardization across data resource. Our assessment calls for efforts to simplify and standardize this information, enabling researchers to more easily evaluate and select appropriate resources when developing DMS Plans or seeking data for secondary analyses. Such improvement would enhance data findability and streamline data sharing in IID research.

## Data Availability

All data generated or analyzed during this study are included in this published article and its supplementary information files, which are available on Figshare.

https://figshare.com/s/ad16d466cc5de818a09e

https://figshare.com/s/d687c78c7eba8b66b056

https://figshare.com/s/892f31b3c663a7bc4cdd

## List of abbreviations

DMS: Data Management and Sharing
IID: Infectious and Immune-mediated Diseases
NIH: National Institutes of Health
NIAID: National Institute of Allergy and Infectious Diseases
FAIR: Findable, Accessible, Interoperable, Reusable
TRUST: Transparency, Responsibility, User Community, Sustainability, and Technology
OSTP: Office of Science and Technology Policy
NLM: National Library of Medicine
MeSH: Medical Subject Headings
DOI: Digital Object Identifier
IRI: Internationalized Resource Identifier
URL: Uniform Resource Locator
dbGaP: Database of Genotypes and Phenotypes
IRB: Institutional Review Board

## Declarations Clinical trial number

Not applicable

## Ethics approval and consent to participate

Not applicable.

## Consent for publication

Not applicable.

## Availability of data and materials

All data generated or analyzed during this study are included in this published article and its supplementary information files, which are available on Figshare at the links provided in the table below.

| Table Number | Description | Link | DOI |
| --- | --- | --- | --- |
| Supplementary Table 1 | Data resources and associated URLs identified from publicly available websites and in consultation with National Institute of Allergy and Infectious Diseases-affiliated subject matter experts (n = 303). | https://figshare.com/s/ad16d466cc5de818a09e | 10.6084/m9.figshare.28832105 |
| Supplementary Table 2 | Main characteristics of reviewed infectious and immune-mediated data resources (n = 58) extended. | https://figshare.com/s/d687c78c7eba8b66b056 | 10.6084/m9.figshare.28843079 |
| Supplementary Table 6 | Assessment of infectious and immune-mediated data resources (n = 19) using a 23-question questionnaire on data submission and resource characteristics. | https://figshare.com/s/892f31b3c663a7bc4cdd | 10.6084/m9.figshare.28843085 |

*Note:* DOIs will go live after embargo.

## Authors’ contributions

Conceptualization: DP, LM; Methodology: DP, LM, RS; Software: DP; LM; Validation: DP, LM, RS; Formal analysis: DP, LM; Investigation: DP, LM; Data curation: DP, LM; Writing – original draft: DP, LM, RS; Writing – review & editing: DP, LM; SF; MH; RS; WVP; Visualization: DP, LM; Supervision: RS; Project administration: RS; Funding acquisition: RS.

## Acknowledgements

Resource recommendations and feedback provided by Scripps, Dr. Maria Giovanni, and NIAID’s Office of Communications and Government Relations.

## Supplement

**Supplementary Table 1.** Data resources and associated URLs identified from publicly available websites and in consultation with National Institute of Allergy and Infectious Diseases-affiliated subject matter experts (n = 303). Available in FigShare: https://figshare.com/s/ad16d466cc5de818a09e

**Supplementary Table 2.** Main characteristics of reviewed infectious and immune-mediated data resources (n = 58) including resource acronyms, if applicable, and URLs. Available in FigShare: https://figshare.com/s/d687c78c7eba8b66b056

**Supplementary Table 3.** Counts by primary diseases or pathogens.

| Category | Count |
| --- | --- |
| General Infectious Diseases and Pathogens | 29 |
| Respiratory Pathogens | 10 |
| HIV/AIDS | 8 |
| Immunological and Autoimmune Diseases | 5 |
| Hemorrhagic Fever Viruses | 3 |
| Arthropod-borne Pathogens | 1 |
| Aspergillosis | 1 |
| Hepatitis C | 1 |
| Papillomaviruses | 1 |
| **Total** | **59\*** |

\*ChemDB HIV, Opportunistic Infection and Tuberculosis Therapeutics Database is included in both the HIV/AIDS category and Respiratory Pathogens.

**Supplementary Table 4.** Counts by data access categories.

| Category | Count |
| --- | --- |
| Open | 34 |
| Controlled | 15 |
| Open; Controlled | 5 |
| Registration | 3 |
| Registration; Controlled | 1 |
| **Total** | **58** |

Data access types were previously defined by Lin *et al.*, 2024. “Open and controlled” indicates that some data are freely accessible without any requirements for the user to register, log in, or pay; some data requires a registration and additional measures to access the data.

**Supplementary Table 5.** Counts by scientific content categories.

| Scientific Content Category | Count |
| --- | --- |
| Omics Data | 38 |
| Clinical Data | 21 |
| Biological Assay Data | 20 |
| Epidemiological Data | 16 |
| Biospecimens | 8 |
| Metadata Catalog | 3 |
| Laboratory Chemicals | 2 |
| Software | 1 |
| Imaging Data | 1 |
| **Total** | **110\*** |

\*Most data resources were characterized under several scientific content categories

