## Supplementary Table 6 for "Infectious, Allergic, and Immune-Mediated Disease Data Resources: A Landscape Overview and Subset Assessment"

**Supplementary Table 6.** Assessment of infectious and immune-mediated data resources (n = 19) using a 23-question questionnaire on data submission and resource characteristics.

| Category | # | Question | Data Resource | | | | | | | | | | | | | | | | | | |
|---|---|---|---|---|---|---|---|---|---|---|---|---|---|---|---|---|---|---|---|---|---|
| | | | **ACDN** | **ClinEpiDB** | **RADx Data Hub** | **CMBTIR** | **dbGaP** | **GISAID** | **HPTN** | **ImmPort** | **IDDO** | **ITN TrialShare** | **MalariGEN** | **mapMECFS** | **N3C** | **Pathoplexus** | **Qiita** | **TB Portals** | **VDJServer** | **USCS Genome** | **VeuPathDB** |
| **1) Access and Submission** | 1.1 | Does the resource accept data submission? | Submission Allowed with Additional Approvals or Contracts | Submission Allowed with Additional Approvals or Contracts | Submission Allowed with Membership | Submission Allowed with Membership | Submission Allowed with Registration or Account | Submission Allowed with Registration or Account | Submission Allowed with Membership | Submission Allowed with Registration or Account | Submission Allowed with Additional Approvals or Contracts | Submission Allowed with Membership | Submission Allowed with Additional Approvals or Contracts | Submission Allowed with Additional Approvals or Contracts | Submission Allowed with Additional Approvals or Contracts | Submission Allowed with Registration or Account | Submission Allowed with Registration or Account | Submission Allowed with Additional Approvals or Contracts | Submission Allowed with Registration or Account | Submission Allowed with Registration or Account | Submission Allowed with Registration or Account |
| | 1.2 | Does the data resource provide open access data? | No | No | No | Yes | Yes | No | No | No | No | No | No | No | Yes | Yes | No | Yes | Yes | Yes | No |
| | 1.3 | Does the data resource require registration (e.g., email) for data access? | No | No | No | No | No | Yes | Yes | Yes | No | Yes | Yes | No | No | No | Yes | No | No | No | Yes |
| | 1.4 | Does the data resource provide controlled access data? | Yes | Yes | Yes | Yes | Yes | No | Yes | Yes | Yes | Yes | Yes | Yes | Yes | No | No | Yes | No | No | No |
| | 1.5 | Does the data resource provide open access metadata? | Yes | Yes | Yes | Yes | Yes | No | Yes | Yes | Yes | No | Yes | No | Yes | Yes | Yes | Yes | Yes | Yes | No |
| | 1.6 | Does the data resource support authentication of data submitters? | Yes | Yes | Yes | Yes | Yes | Yes | Yes | Yes | Yes | Yes | Yes | Yes | Yes | Yes | Yes | Yes | Yes | Yes | Yes |
| | 1.7 | Does the data resource have formatting requirement for data submission? | No | No | No | Yes | Yes | No | No | Yes | No | No | No | Yes | Yes | Yes | Yes | No | No | Yes | Yes |
| | 1.8 | Does the data resource have size limit requirements for data submission? | No | No | No | No | No | No | Yes | No | No | No | No | No | No | No | No | No | No | No | No |
| | 1.9 | Are there costs associated with depositing the data? | No | No | No | No | No | No | No | No | No | No | No | No | No | No | No | No | No | No | Yes* |
| **2) Identification, Provenance, and Quality Assurance** | 2.1 | Does the data resource assign each dataset an identifier? If yes, is it a persistent or internal identifier? | No | Persistent Identifier | Persistent Identifier | Internal Identifier | Internal Identifier | Internal Identifier | Persistent Identifier | Persistent Identifier | Persistent Identifier | Internal Identifier | Internal Identifier | No | No | Internal Identifier | Internal Identifier | Internal Identifier | Internal Identifier | Internal Identifier | Internal Identifier |
| | 2.2 | Does the data resource have a system in place to track provenance to the (meta)data? | Yes | Yes | Yes | No | Yes | Yes | Yes | Yes | No | Yes | No | Yes | Yes | Yes | No | No | Yes | Yes | Yes |
| | 2.3 | Does the data resource support | No | No | Yes | Yes | Yes | Yes | Yes | Yes | Yes | No | Yes | Yes | Yes | Yes | No | Yes | No | No | Yes |

| | | | | | | | | | | | | | | | | | | | | | |
|---|---|---|---|---|---|---|---|---|---|---|---|---|---|---|---|---|---|---|---|---|---|
| | | expert curation or quality assurance to improve the accuracy and integrity of datasets and metadata? | | | | | | | | | | | | | | | | | | | |
| **3) Data Retrieval and Tools** | 3.1 | Can the (meta)data be accessed through an API? | No | No | No | No | Yes | Yes | Yes | Yes | No | No | Yes | Yes | No | Yes | Yes | Yes | Yes | Yes | Yes |
| | 3.2 | Can the user download the data to their local machine? | Yes | Yes | Yes | Yes | Yes | Yes | Yes | Yes | Yes | Yes | Yes | Yes | Yes | Yes | Yes | Yes | Yes | Yes | Yes |
| | 3.3 | Does the data resource provide data analytical tools? | No | No | Yes | Yes | No | Yes | No | Yes | No | Yes | Yes | Yes | Yes | No | Yes | Yes | Yes | Yes | Yes |
| | 3.4 | Does the data resource provide a workspace? | No | No | Yes | Yes | No | No | No | Yes | No | Yes | Yes | No | Yes | No | Yes | Yes | No | No | Yes |
| | 3.5 | Are these costs associated with maintaining data in the workspace? | NA | NA | No | No | NA | NA | NA | No | NA | No | No | NA | No | NA | No | No | NA | NA | Yes |
| | 3.6 | Are users able to utilize their own analytical tools within the workspace? | NA | NA | No | No | NA | NA | NA | No | NA | No | No | NA | Yes | NA | Yes | No | NA | NA | No |
| | 3.7 | Are there costs associated with analyzing the data in the workspace? | NA | NA | No | No | NA | NA | NA | No | NA | No | No | NA | No | NA | No | No | NA | NA | No |
| **4) Documentation and Compliance** | 4.1 | Does the data resource provide documentation on risk management (e.g., data breach, natural disasters)? | No | No | No | Yes | Yes | No | Yes | Yes | Yes | No | No | No | Yes | Yes | Yes | Yes | No | No | No |
| | 4.2 | Does the data resource provide documentation on its data retention policies? | No | No | No | No | No | Yes | Yes | No | Yes | No | No | No | No | Yes | No | No | No | No | No |
| | 4.3 | Does the data resource have security policies in place that ensure protection against unauthorized access, modification, or release of data, with appropriate | No | No | Yes | Yes | Yes | Yes | Yes | Yes | Yes | Yes | Yes | Yes | Yes | Yes | Yes | Yes | Yes | No | Yes |

| | | | | | | | | | | | | | | | | | | | | | |
|---|---|---|---|---|---|---|---|---|---|---|---|---|---|---|---|---|---|---|---|---|---|
| | | security levels based on data sensitivity? | | | | | | | | | | | | | | | | | | | |
| | 4.4 | Does the data resource provide documentation for its terms for data use? | Yes | Yes | Yes | Yes | Yes | Yes | Yes | Yes | Yes | Yes | Yes | Yes | Yes | Yes | Yes | Yes | No | Yes | Yes |

*Abbreviations: ACDN, AccessClinicalData@NIAID; ClinEpiDB, Clinical Epidemiology Database; RADx Data Hub, COVID RADx Data Hub; CMBTIR, Center for International Blood & Marrow Transplant Research; dbGaP, Database of Genotypes and Phenotypes; GISAID, Global Initiative on Sharing All Influenza Data; HPTN, HIV Prevention Trials Network; ImmPort, Immunology Database and Analysis Portal; IDDO, Infectious Diseases Data Observatory; ITN TrialShare, Immune Tolerance Network TrialShare; MalariaGEN, Malaria Genomic Epidemiology Network; mapMECFS, Myalgic Encephalomyelitis/Chronic Fatigue Syndrome Data Platform; N3C, National COVID Cohort Collaborative; TB Portals, Tuberculosis Data Portals; UCSC Genome Browser, University of Santa Cruz Genome Browser; VEuPathDB, Eukaryotic Pathogen Database Resources.* This assessment was conducted by NIAID based on publicly available information. If you notice any discrepancies, please contact the authors.
