## Supplementary Table 2 for "Infectious, Allergic, and Immune-Mediated Disease Data Resources: A Landscape Overview and Subset Assessment"

**Supplementary Table 2.** Main characteristics of reviewed infectious and immune-mediated data resources (n = 58) including resource acronyms, if applicable, and URLs.

| Data Resource Name | Data Resource Name Acronym, if Applicable | URL | Primary Disease or Pathogen | Scientific Content | Data Access | Data Accepted |
|---|---|---|---|---|---|---|
| AccessClinicalData@NIAID | ACDN | https://accessclinicaldata.niaid.nih.gov/ | Respiratory Pathogens | Biological Assay; Clinical; Epidemiological | Controlled | Yes |
| ACTG/IMPAACT Specimen Repository | ACTG/IMPAACT | https://www.impaactnetwork.org/resources/lab-center/specimen-repository | HIV/AIDS | Biospecimens; Clinical | Controlled | No |
| Aspergillus Genome Database | AspGD | https://mycocosm.jgi.doe.gov/Aspnid1/Aspnid1.home.html | Aspergillosis | Omics | Open | No |
| BacDive | | https://bacdive.dsmz.de/ | General Infectious Diseases and Pathogens | Biological Assay | Open | No |
| Bacterial and Viral Bioinformatics Resource Center | BV-BRC | https://www.bv-brc.org/ | General Infectious Diseases and Pathogens | Omics | Open | No |
| BEIResources | BEI | https://www.beiresources.org/ | General Infectious Diseases and Pathogens | Laboratory Chemicals | Controlled; Open | No |
| BioCyc | | https://biocyc.org/ | General Infectious Diseases and Pathogens | Omics | Open | No |
| Biological General Repository for Interaction Datasets | BioGRID | https://thebiogrid.org/ | General Infectious Diseases and Pathogens | Omics | Open | No |
| Center for International Blood & | CIBMTR | https://www.cibmtr.org/ | Immunological and Autoimmune Diseases | Biospecimens; Clinical | Controlled; Open | Yes |

| | | | | | | |
|---|---|---|---|---|---|---|
| Marrow Transplant Research | | | | | | |
| Center for Viral Systems Biology | CVSB | https://cvisb.org/ | Hemorrhagic Fever Viruses | Biological Assay; Biospecimens; Clinical; Epidemiological; Omics | Open | No |
| ChemDB HIV, Opportunistic Infection and Tuberculosis Therapeutics Database | ChemDB | https://chemdb.niaid.nih.gov/ | HIV/AIDS; Respiratory Pathogens | Laboratory Chemicals | Open | No |
| ClinEpiDB | | https://clinepidb.org/ | General Infectious Diseases and Pathogens | Clinical; Epidemiological | Controlled; Open | Yes |
| COVID-19 Research Database | COVID-19 RDB | https://covid19researchdatabase.org/ | Respiratory Pathogens | Clinical; Epidemiological; Omics | Controlled | No |
| COVID RADx Data Hub | RADx Data Hub | https://radxdatahub.nih.gov/ | Respiratory Pathogens | Biological Assay; Clinical; Epidemiological; Omics | Controlled | Yes |
| Database of Antimicrobial Activity and Structure of Peptides | DBAASP | https://dbaasp.org/home | General Infectious Diseases and Pathogens | Omics | Open | No |
| Database of Genotypes and Phenotypes | dbGaP | https://www.ncbi.nlm.nih.gov/gap/ | General Infectious Diseases and Pathogens | Omics | Controlled | Yes |
| Data Discovery Engine-registered Datasets | DDE | https://discovery.biothings.io/ | General Infectious Diseases and Pathogens | Metadata Catalog | Open | No |

| | | | | | | |
|---|---|---|---|---|---|---|
| The Global Health Observatory | GHO | https://www.who.int/data/gho | General Infectious Diseases and Pathogens | Epidemiological | Open | No |
| Global Initiation Sharing All Influenza Data | GISAID | https://gisaid.org/ | Respiratory Pathogens | Clinical; Epidemiological; Omics | Registration | Yes |
| Hemorrhagic Fever Viruses Database Project | HFVDB | https://hfv.lanl.gov/content/index/ | Hemorrhagic Fever Viruses | Biological Assay; Omics | Open | No |
| Hepatitis C Virus Database Project | HCVDB | https://hcv.lanl.gov/content/index | Hepatitis C | Biological Assay; Omics | Open | No |
| Heterogeneity in Human Immune Cells | HIC | https://heterogeneity.niaid.nih.gov/ | General Infectious Diseases and Pathogens | Biological Assay | Open | No |
| HIV Databases | HIVDB | https://www.hiv.lanl.gov/content/index | HIV/AIDS | Omics; Biological Assay | Open | No |
| HIV Prevention Trials Network | HPTN | https://www.hptn.org/ | HIV/AIDS | Biological Assay; Biospecimens; Clinical; Epidemiological; Omics | Controlled; Open | Yes |
| HIV Vaccine Trials Network | HVTN | https://www.hvtn.org/ | HIV/AIDS | Biospecimens; Clinical | Controlled | No |
| Human Microbiome Project Portal | HMP | https://portal.hmpdacc.org/ | General Infectious Diseases and Pathogens | Omics | Open | No |
| ImmPort | | https://www.immport.org/shared/home | General Infectious Diseases and Pathogens | Biological Assay; Clinical; Omics | Controlled; Registration | Yes |
| Immune Epitope Database | IEDB | https://www.iedb.org/ | General Infectious Diseases and Pathogens | Biological Assay | Open | No |
| ImmuneSpace | | https://immunespace.org/ | General Infectious Diseases and Pathogens | Biological Assay; Omics | Open | No |

| | | | | | | |
|---|---|---|---|---|---|---|
| Immune Tolerance Network TrialShare | ITN TrialShare | https://www.immunetolerance.org/for-researchers/trialshare | General Infectious Diseases and Pathogens | Biospecimens; Clinical | Controlled | No |
| Immunological Genome Project | ImmGen | https://www.immgen.org/ | General Infectious Diseases and Pathogens | Biological Assay; Omics | Open | No |
| Infectious Diseases Data Observatory | IDDO | https://www.iddo.org/ | General Infectious Diseases and Pathogens | Clinical; Epidemiological; Omics | Controlled | Yes |
| The Institute for Genome Sciences at the University of Maryland School of Medicine Genomic Center for Infectious Diseases | IGS | https://gcid.igs.umaryland.edu/ | General Infectious Diseases and Pathogens | Omics | Open | No |
| International Committee Taxonomy of Viruses | ICTV | https://ictv.global/ | General Infectious Diseases and Pathogens | Metadata Catalog | Open | No |
| iReceptor | | https://gateway.ireceptor.org/login | General Infectious Diseases and Pathogens | Biological Assay | Registration | No |
| MACS/WIHS Combined Cohort Study | MWCCS | https://statepi.jhsph.edu/mwccs/ | HIV/AIDS | Biospecimens; Clinical; Epidemiological | Controlled | No |
| Malaria Genomic Epidemiology Network | MalariaGEN | https://www.malariagen.net/ | Arboviruses | Epidemiological; Omics | Controlled; Open | Yes |
| mapMECFS | | https://mapmecfs.org/ | Immunological and Autoimmune Diseases | Omics; Epidemiological; Biological Assay | Controlled | Yes |
| MTB Network Portal | MTB Portal | https://networks.systemsbiology.net/mtb/ | Respiratory Pathogens | Omics; Software | Open | No |

| | | | | | | |
|---|---|---|---|---|---|---|
| Microbicide Trials Network | MTN | https://www.mtnstopshiv.org/ | HIV/AIDS | Clinical | Controlled | No |
| Mycobrowser | | https://mycobrowser.epfl.ch/ | Respiratory Pathogens | Omics | Open | No |
| National Center for Advancing Translational Sciences OpenData Portal | OpenData Portal | https://opendata.ncats.nih.gov/covid19 | Respiratory Pathogens | Biological Assay; Clinical | Open | No |
| National Center for Biotechnology Information Virus | NCBI Virus | https://www.ncbi.nlm.nih.gov/labs/virus/vssi/#/ | General Infectious Diseases and Pathogens | Omics | Open | Yes |
| National COVID Cohort Collaborative | N3C | https://covid.cd2h.org/ | Respiratory Pathogens | Biological Assay; Clinical; Epidemiological | Controlled | Yes |
| Open Germline Receptor Database | OGRDB | Open Germline Receptor Database | Immunological and Autoimmune Diseases | Omics | Open | No |
| Papillomavirus Episteme | PaVE | https://pave.niaid.nih.gov/ | Papillomaviruses | Omics | Open | No |
| Pathoplexus | | https://pathoplexus.org/ | Hemorrhagic Fever Viruses | Omics | Open | Yes |
| Project TYCHO | TYCHO | https://www.tycho.pitt.edu/ | General Infectious Diseases and Pathogens | Epidemiological | Open | No |
| Qiita | | https://qiita.ucsd.edu/ | General Infectious Diseases and Pathogens | Omics | Registration | Yes |
| Stanford University HIV Drug Resistance Database | Stanford HIV DB | https://hivdb.stanford.edu/ | HIV/AIDS | Biological Assay; Clinical; Omics | Open | No |

| | | | | | | |
|---|---|---|---|---|---|---|
| Structural Database of Allergenic Proteins | SDAP | https://fermi.utmb.edu/ | Immunological and Autoimmune Diseases | Omics | Open | Yes |
| TB Portals | | https://tbportals.niaid.nih.gov/ | Respiratory Pathogens | Clinical; Epidemiological; Imaging; Omics | Controlled | Yes |
| United States Immunodeficiency Network | USIDNET | https://usidnet.org/ | Immunological and Autoimmune Diseases | Biospecimens; Clinical; Omics | Controlled | No |
| University of Santa Cruz Genome Browser | UCSC Genome Browser | https://genome.ucsc.edu/ | General Infectious Diseases and Pathogens | Omics | Open | Yes |
| Vaccine Investigation and Online Information Network | VIOLIN | https://violinet.org/ | General Infectious Diseases and Pathogens | Biological Assay; Metadata Catalog; Omics | Open | No |
| VDJbase | | https://vdjbase.org/ | General Infectious Diseases and Pathogens | Omics | Open | No |
| VDJServer | | https://vdjserver.org/ | General Infectious Diseases and Pathogens | Omics | Open | Yes |
| VEuPathDB | | https://veupathdb.org/veupathdb/app | General Infectious Diseases and Pathogens | Biological Assay; Clinical; Epidemiological; Omics | Open | No |

Characteristics for each data resource including the resource name, primary disease or pathogen and scientific content (i.e., categories or types) of hosted data, data access status (either open, controlled, or registration indicating that registering an account with the data resource is necessary to view the data), and indication whether data can be deposited. *Abbreviations: ACTG, AIDS Clinical Trials Group; IMPAACT, International Maternal Pediatric Adolescent AIDS Clinical Trial Network; HIV, Human Immunodeficiency Virus; MACS, Multicenter AIDS Cohort Study; WIHS, Women's Interagency*

*HIV Study; TB, Tuberculosis*. Primary Infectious Disease: General Infectious Diseases and Pathogens indicates data resources include data relevant to various diseases and conditions, including infectious and immune-mediated disease data.
