## Supplementary Table 1 for "Infectious, Allergic, and Immune-Mediated Disease Data Resources: A Landscape Overview and Subset Assessment"

**Supplementary Table 1.** Data resources and associated URLs identified from publicly available websites and in consultation with National Institute of Allergy and Infectious Diseases-affiliated subject matter experts (n = 303).

| Name | URL |
|---|---|
| 1000 Genomes Project | https://www.internationalgenome.org/data |
| 3D Reconstruction via Stereoscopy for the Study of Mosquito Swarms | https://www.niaid.nih.gov/research/3d-reconstruction-stereoscopy-study-mosquito-swarms |
| 4D Nucleome (4DN) | https://www.4dnucleome.org/ |
| AccessClinicalData@NIAID | https://www.niaid.nih.gov/research/access-clinical-data-niaid |
| ACTG/IMPAACT Specimen Repository | http://www.specimenrepository.org/RepositorySite/home.html |
| AddGene | https://www.addgene.org/ |
| AIDS Clinical Trials Group (ACTG) | https://actgnetwork.org/ |
| ALEdb | https://aledb.org/ |
| Allele Frequency Net Database | http://www.allelefrequencies.net/ |
| Allen Institute Human Immune System Explorer | https://explore.allenimmunology.org/ |
| Alliance of Genome Resources | https://www.alliancegenome.org/ |
| AllofUs Research Program | https://allofus.nih.gov/ |
| AmoebaDB | https://amoebadb.org/amoeba/app |
| AntibodyRegistry | https://www.antibodyregistry.org/ |
| ArrayExpress | https://www.ebi.ac.uk/biostudies/arrayexpress |
| Aspergillus Genome Database | https://mycocosm.jgi.doe.gov/Aspnid1/Aspnid1.home.html |
| aspredicted.org | https://aspredicted.org/ |
| BacDive | https://bacdive.dsmz.de/ |
| Bacterial and Viral Bioinformatics Resource Center (BV-BRC) | https://www.bv-brc.org/ |
| BEIResources | https://www.beiresources.org/ |
| BibTri v3.0 | https://bibtri.cepave.edu.ar/webbibtri.php?lang=en |
| bio.tools | https://bio.tools/ |
| Bioconductor | https://www.bioconductor.org/ |
| BioContainers | https://biocontainers.pro/ |
| Biocyc | https://biocyc.org/ |
| Bioinformatics and Computational Biosciences Branch Services | https://www.niaid.nih.gov/research/bcbb-services |
| Biological General Repository for Interaction Datasets (BioGRID) | https://thebiogrid.org/ |
| BioModels | https://www.ebi.ac.uk/biomodels/ |
| Bioregistry | https://bioregistry.io/ |
| CardioVascular Research Grid | hthttps://www.cvrgrid.org/ |
| Cell Collective | https://cellcollective.org/# |

| | |
|---|---|
| Cell Image Library | http://www.cellimagelibrary.org/ |
| Cellosaurus | https://www.cellosaurus.org/ |
| Center for International Blood & Marrow Transplant Research | https://www.cibmtr.org/Pages/index.aspx |
| Center for Open Science Pre-registration | https://www.cos.io/initiatives/prereg |
| Center for Viral Systems Biology (CViSB) | https://cvisb.org/ |
| Centers for Research in Emerging Infectious Diseases (CREID) | https://creid-network.org/ |
| Centers of Excellence for Influenza Research and Response (CEIRR) | https://www.ceirr-network.org/ |
| ChemDB HIV, Opportunistic Infection and Tuberculosis Therapeutics Database | https://chemdb.niaid.nih.gov/ |
| ChemokineDB | https://bioinformatics.niaid.nih.gov/chemokinedb/ |
| Chicago Center for Functional Annotation (CCFA) | https://www.bv-brc.org/webpage/website/data_collections/content/ccfa.html |
| CiteAb | https://www.citeab.com/ |
| ClinEpiDB | https://clinepidb.org/ce/app |
| ClinicalGenomeResource (ClinGen) | https://www.clinicalgenome.org/ |
| ClinicalTrials.gov | https://clinicaltrials.gov/ |
| ClinRegs | https://clinregs.niaid.nih.gov/ |
| ClinVar | https://www.ncbi.nlm.nih.gov/clinvar/intro/ |
| Code Ocean | https://codeocean.com/ |
| Columbia Lyme Disease Specimen Bank | https://www.columbia-lyme.org/columbia-specimen-bank |
| Comprehensive R Archive Network | https://cran.r-project.org/ |
| Cooperative Centers on Human Immunology (CCHI) | https://www.niaid.nih.gov/research/cooperative-centers-human-immunology |
| Coronavirus Resources for Researchers | https://www.niaid.nih.gov/diseases-conditions/coronavirus-resources |
| COVID RADx Data Hub | https://radx-hub.nih.gov/home |
| COVID-19 Research Database | https://covid19researchdatabase.org/ |
| CryptoDB | https://cryptodb.org/cryptodb/app |
| DAIDS Regulatory Support Center (RSC) | https://www.niaid.nih.gov/research/daids-regulatory-support-center |
| Data and Specimen Hub (DASH) | https://dash.nichd.nih.gov/ |
| Data Discovery Engine-registered Datasets | https://discovery.biothings.io/dataset?guide=/guide/niaid |
| Database of Antimicrobial Activity and Structure of Peptides (DBAASP) | https://dbaasp.org/home |

| | |
|---|---|
| Database of Genotypes and Phenotypes (dbGaP) | https://www.ncbi.nlm.nih.gov/gap/ |
| Database of Mutations Causing Human Hyper IgE Syndrome (STAT3base) | https://www.niaid.nih.gov/research/stat3base |
| Database of Short Genetic Variations (dbSNP) | https://www.ncbi.nlm.nih.gov/snp/ |
| DisProt database | https://disprot.org/ |
| Distributed Archives for Neurophysiology Data Integration (DANDI) | https://dandiarchive.org/ |
| Dockstore | https://dockstore.org/ |
| Dryad | https://datadryad.org/ |
| EcoCyc | https://ecocyc.org/ |
| eDGAR | http://edgar.biocomp.unibo.it/gene_disease_db/ |
| Electron Microscopy Data Bank (EMDB) | https://www.ebi.ac.uk/emdb/ |
| ENCODE Project | https://www.encodeproject.org/about/data-access/ |
| Ensembl | https://useast.ensembl.org/index.html |
| European Genome-Phenome Archive (EGA) | https://ega-archive.org/ |
| European Nucleotide Archive (ENA) | https://www.ebi.ac.uk/ena/browser/ |
| European Variation Archive (EVA) | https://www.ebi.ac.uk/eva/ |
| ExpressionAtlas | https://www.ebi.ac.uk/gxa/home |
| Extracellular RNA Communication (ExRNA) Atlas | https://exrna-atlas.org/ |
| FAIRDOMHub | https://fairdomhub.org/ |
| figshare | https://figshare.com/ |
| Filariasis Research Reagent Resource Center (FR3) | http://www.filariasiscenter.org/ |
| FlowRepository | https://flowrepository.org/public_experiment_representations |
| Flu Hub | https://www.fluhub.org/ |
| FlyBase | https://flybase.org/ |
| Functional Lists of Unknown TB Entities (FLUTE) | in bv brc |
| FungiDB | https://fungidb.org/fungidb/app |
| Gemma | https://gemma.msl.ubc.ca/expressionExperiment/showAllExpressionExperiments.html |
| GenBank | https://www.ncbi.nlm.nih.gov/genbank/ |
| Gencode | https://www.gencodegenes.org/ |
| Gene Expression Omnibus (GEO) | https://www.ncbi.nlm.nih.gov/geo/ |
| Gene Ontology | http://geneontology.org/ |
| GeneNetwork | https://uswest.ensembl.org/index.html |

| | |
|---|---|
| Generalized Proteomics Data Meta-analysis Database (GPMDB) | https://gpmdb.thegpm.org/ |
| Genes Unknown in Acinetobacter baumannii (GUNK) | https://www.bv-brc.org/webpage/website/data_collections/content/gunk.html |
| GenitoUrinary Development Molecular Anatomy Project (GUDMAP) | https://www.atlas-d2k.org/gudmap/ |
| Genomic Centers for Infectious Diseases (GCID) Resources | https://www.niaid.nih.gov/research/gcid-resources |
| Genotype-Tissue Expression (GTEx) Portal | https://gtexportal.org/home |
| GiardiaDB | https://giardiadb.org/giardiadb/app |
| GISAID | https://gisaid.org/ |
| GitHub | https://github.com/ |
| Global Natural Products Social Molecular Networking (GNPS) | https://gnps.ucsd.edu/ProteoSAFe/static/gnps-splash.jsp |
| Global Vector Hub | https://globalvectorhub.lshtm.ac.uk/ |
| GlyGen | https://data.glygen.org/ |
| gnomAD | https://gnomad.broadinstitute.org/ |
| GWAS Catalog | https://www.ebi.ac.uk/gwas/ |
| H3Africa | https://www.h3abionet.org/resources/h3africa-archive |
| Harvard DataVerse | https://dataverse.harvard.edu/ |
| Hemorrhagic Fever Viruses (HFV) Database Project | https://hfv.lanl.gov/content/index |
| Hepatitis C Virus Database Project (HCV) | https://hcv.lanl.gov/content/index |
| Heterogeneity in Human Immune Cells | https://heterogeneity.niaid.nih.gov/ |
| HIV Databases | https://www.hiv.lanl.gov/content/index |
| HIV Prevention Trials Network (HPTN) | https://www.hptn.org/ |
| HIV Vaccine Trials Network (HVTN) | https://www.hvtn.org/ |
| HostDB | https://hostdb.org/hostdb/app |
| HuBMap | https://hubmapconsortium.org/ |
| HUGO Gene Nomenclature Committee (HGNC) | https://www.genenames.org/ |
| Human Cell Atlas | https://www.humancellatlas.org/ |
| Human Immunology Project Consortium | https://www.immuneprofiling.org/hipc/page/show |
| Human Microbiome Project Portal | https://portal.hmpdacc.org/projects/t |
| ICGC Data Portal | https://dcc.icgc.org/ |
| Illuminating the Drugable Genome (IDG) | https://druggablegenome.net/ |
| Immcantation Portal | https://immcantation.readthedocs.io/en/stable/ |

| | |
|---|---|
| ImmPort | https://www.immport.org/shared/home |
| Immune Epitope Database (IEDB) | https://www.iedb.org/ |
| immuneAccess | https://clients.adaptivebiotech.com/immuneaccess |
| ImmuneSpace | https://immunespace.org/ |
| ImmuneXpresso | http://immuneexpresso.org/immport-immunexpresso/public/immunexpresso/search |
| Immuno Polymorphism Database (IPD) | https://www.ebi.ac.uk/ipd/ |
| Immunological Genome Project | https://www.immgen.org/ |
| Immunophenotyping Assessment in COVID-19 Cohort (IMPACC) | https://docs.immport.org/home/impaccslides/ |
| iModulonDB | https://imodulondb.org/ |
| INCLUDE Data Coordination Center | https://includedcc.org/ |
| IndraDB | https://db.indra.bio/ |
| Infectious Diseases Data Observatory (IDDO) | https://www.iddo.org/ |
| Influenza Research Database | https://www.fludb.org/ |
| innateDB | https://www.innatedb.com/ |
| Integrated Analysis Of Multimodal Single-Cell Data | https://atlas.fredhutch.org/nygc/multimodal-pbmc/ |
| Integrated Human Microbiome Project (iHMP) | https://hmpdacc.org/ihmp/ |
| International Committee Taxonomy of Viruses (ICTV) | https://ictv.global/ |
| International epidemiology databases to evaluate AIDS (IeDEA) | https://www.iedea.org/ |
| International Human Epigenome Consortium (IHEC) | https://epigenomesportal.ca/ihec/ |
| International Maternal Pediatric Adolescent AIDS Clinical Trials network (IMPAACT) | https://www.impaactnetwork.org/ |
| InterPro | https://www.ebi.ac.uk/interpro/ |
| IPD-IMGT/HLA | https://www.ebi.ac.uk/ipd/imgt/hla/ |
| iReceptor | https://gateway.ireceptor.org/login |
| ITN TrialShare | https://www.immunetolerance.org/researchers/trialshare |
| JGI Genome Portal | https://genome.jgi.doe.gov/portal/ |
| JPOST Repository | https://globe.jpostdb.org/ |
| Kids First | https://www.notion.so/Studies-and-Access-a5d2f55a8b40461eac5bf32d9483e90f |
| Knockout Mouse Phenotyping Program (KOMP2) | https://www.mousephenotype.org/ |
| Library of Integrated Network-based Cellular Signatures (LINCS) | https://lincsproject.org/ |

| | |
|---|---|
| MACS/WIHS Combined Cohort | https://statepi.jhsph.edu/mwccs/ |
| Malaria Genomic Epidemiology Network (MalariaGEN) | https://www.malariagen.net/ |
| MassBank Database | https://massbank.eu/MassBank/Search |
| MassIVE | https://massive.ucsd.edu/ProteoSAFe/static/massive.jsp |
| Mendeley Data | https://data.mendeley.com/ |
| MetaboLights | https://www.ebi.ac.uk/metabolights/ |
| MetabolomeExpress | http://www.metabolome-express.org/ |
| Metabolomics Workbench | https://www.metabolomicsworkbench.org/ |
| Microbicide Trials Network (MTN) | https://www.mtnstopshiv.org/ |
| MicrobiomeDB | https://microbiomedb.org/mbio/app/ |
| MicrosporidiaDB | https://microsporidiadb.org/micro/app |
| Molecular Transducers of Physical Activity in Humans (MoTrPAC) | https://motrpac-data.org/ |
| Mouse Genome Informatics | https://www.informatics.jax.org/ |
| Mouse Organogenesis Cell Atlas | https://oncoscape.v3.sttrcancer.org/atlas.gs.washington.mouse.rna/downloads |
| Mouse Phenome Database | https://phenome.jax.org/ |
| MTB Network Portal | http://networks.systemsbiology.net/mtb/ |
| Multicenter AIDS Cohort Study (MACS) Public Data Set | https://statepi.jhsph.edu/macs/pdt.html |
| Mycobrowser | https://mycobrowser.epfl.ch/ |
| MyExperiment.org | https://myexperiment.org/home |
| NIDA National Addiction & HIV Data Archive Program | https://www.icpsr.umich.edu/web/pages/NAHDAP/index.html |
| National COVID Cohort Collaborative (N3C) | https://covid.cd2h.org/ |
| National Database for Autism Research | https://nda.nih.gov/ |
| National Disease Research Interchange | https://ndriresource.org/ |
| NCBI BioProject | https://www.ncbi.nlm.nih.gov/bioproject |
| NCBI BioSample | https://www.ncbi.nlm.nih.gov/biosample/ |
| NCBI Geo | https://www.ncbi.nlm.nih.gov/geo/ |
| NCBI Sequence Read Archive (SRA) | https://www.ncbi.nlm.nih.gov/sra |
| NCBI Virus | https://www.ncbi.nlm.nih.gov/labs/virus/vssi/#/ |
| NCI Genomic Data Commons (GDC) | https://gdc.cancer.gov/ |
| Nematode.net | Nematode.net |
| Nephele | https://nephele.niaid.nih.gov/ |
| NeuroMorpho.org | NeuroMorpho.org |
| NeuroVault | https://neurovault.org/ |
| NHLBIConnects | https://nhlbi-connects.org/data-request |

| | |
|---|---|
| NIAID Bioinformatics Portal | https://bioinformatics.niaid.nih.gov/ |
| NICHD DASH | http://dash.nichd.nih.gov/ |
| NIDKK Central Repository | https://repository.niddk.nih.gov/home/ |
| NIH AIDS Reagent Program; will be deprecated and combined with BEI Resources on January 14, 2024 | https://www.hivreagentprogram.org/ |
| NIH BioWulf | https://hpc.nih.gov/ |
| NIH CDE Repository | https://cde.nlm.nih.gov/home |
| NIH Common Fund Data Ecosystem | https://app.nih-cfde.org/ |
| NIH RePORTER | https://reporter.nih.gov/ |
| NIHFigShare | https://nih.figshare.com/ |
| NITRC Neuroimaging Data Repository | https://www.nitrc.org/xnat/ |
| NLM Data Discovery | https://datadiscovery.nlm.nih.gov/ |
| NODE | https://www.biosino.org/node/ |
| Nonhuman Primate Radiation Survivor Late Effects Cohort (NHP RSC) | https://www.niaid.nih.gov/research/nonhuman-primate-radiation-survivor-late-effects-cohort |
| Nonhuman Primate Reagent Resource | https://www.nhpreagents.org/ |
| Non-Obese Diabetic (NOD) Mouse BAC Library | https://www.sanger.ac.uk/collaboration/sequencing-of-idd-regions-in-the-nod-mouse-genome/ |
| NYU Data Catalog | https://datacatalog.med.nyu.edu/ |
| Observed Antibody Space | https://opig.stats.ox.ac.uk/webapps/oas/ |
| Omics DI | https://www.omicsdi.org/ |
| Open Provenance Model for Workflows Repository | https://www.opmw.org/ |
| Open Science Framework | https://osf.io/ |
| OpenfMRI.org | https://openfmri.org/ |
| OpenNeuro | https://openneuro.org/ |
| Orfeome | https://www.niaid.nih.gov/research/orfeome |
| ORGDB | https://ogrdb.airr-community.org/ |
| OrthoMCL | https://orthomcl.org/orthomcl/app |
| Panther Classification System | http://pantherdb.org/ |
| Papillomavirus Episteme (PaVE) | https://pave.niaid.nih.gov/ |
| Pathosystems Resource Integration Center (PATRIC) | in bv brc |
| Patient-Reported Outcomes Measurement Information System (PROMIS) | https://www.healthmeasures.net/resource-center/research-tools/datasets-for-your-research |
| PAXDB | https://pax-db.org/ |
| PeptideAtlas | http://www.peptideatlas.org/ |
| PharmGKB | https://www.pharmgkb.org/ |

| | |
|---|---|
| PhenoDB | https://phenodb.org/about |
| PhysioBank | https://archive.physionet.org/physiobank/ |
| Physiome Model Repo | https://models.physiomeproject.org/welcome |
| PiroplasmaDB | https://piroplasmadb.org/piro/app |
| PlasmoDB | https://plasmodb.org/plasmo/app |
| Polygenic Score Catalog | https://www.pgscatalog.org/ |
| Predictive Oncology Model and Data Clearinghouse (MoDaC) | https://modac.cancer.gov/ |
| Primary Immunodeficiency (PI) Diseases Registry | https://www.niaid.nih.gov/research/primary-immunodeficiency-diseases-registry |
| Project TYCHO | https://www.tycho.pitt.edu/ |
| Protein Data Bank | https://www.rcsb.org/ |
| Proteome Xchange | https://www.proteomexchange.org/ |
| Proteomics Identifications Database (PRIDE) | https://www.ebi.ac.uk/pride/markdownpage/searchpridearchive |
| Public Health Image Library | https://phil.cdc.gov/ |
| PubMed Central Code Availability Statements | NA |
| PubMed Central Data Availability Statements | NA |
| PubMed Central Supplemental Information | NA |
| Python Package Index | https://pypi.org/ |
| Qiita | https://qiita.ucsd.edu/ |
| Quantitative Set Analysis for Gene Expression (QuSAGE) | https://www.niaid.nih.gov/research/qusage |
| Rakai Community Cohort Study (RCCS) | https://www.rhsp.org/research/rccs/rccs-overview |
| Rat Genome Database | https://rgd.mcw.edu/ |
| Reactome | https://reactome.org/ |
| reframeDB | https://reframedb.org/ |
| Regional Prospective Observational Research in Tuberculosis (RePORT) | https://reportinternational.org/ |
| Rep-seq dataset Analysis Platform with an Integrated Antibody Database (RAPID) | NA |
| Roadmap Epigenomics Project | https://www.ncbi.nlm.nih.gov/geo/roadmap/epigenomics/ |
| RRID Portal | https://scicrunch.org/resources |
| Saccharomyces Genome Database (SGD) | https://yeastgenome.org/ |
| Scripps Consortium for HIV/AIDS Vaccine Development (CHAVD) | https://www.scripps.edu/science-and-medicine/centers-and-institutes/consortium-for-hiv-aids-vaccine-development/#:~:text=The%20mission%20of%20the%20Consort |

| | |
|---|---|
| | ium,%2C%20genomics%2C%20bioinformatics%20and%20proteomics. |
| Seven Bridges Public Apps Gallery | NA |
| Signaling Pathways Project (SPP) | http://www.signalingpathways.org/index.jsf |
| SimTK | https://simtk.org/ |
| Stanford University HIV Drug Resistance Database (HIVDB) | https://hivdb.stanford.edu/ |
| Stimulating Peripheral Activity to Relieve Conditions (SPARC) | https://sparc.science/about |
| Structural Database of Allergenic Proteins (SDAP) | https://fermi.utmb.edu/ |
| Structural Genomics Centers for Infectious Diseases: Resources | https://www.niaid.nih.gov/research/structural-genomics-centers-infectious-diseases-resources |
| Synapse | https://www.synapse.org/ |
| Systems Biology Consortium Resources | https://www.niaid.nih.gov/research/systems-biology-consortium-resources |
| TB Portals | https://tbportals.niaid.nih.gov/ |
| Texas Medical Center Genomic Center for Infectious Diseases (GCID) | https://gcid.research.bcm.edu/overview |
| The Broad Institute's Genomic Center for Infectious Diseases (GCID) | https://www.broadinstitute.org/scientific-community/science/projects/gscid/genomic-center-infectious-diseases |
| The Cancer Genome Characterization Initiative (CGCI) | NA |
| The Cancer Imaging Archive (TCIA) | NA |
| The Dataverse Project | https://dataverse.org/ |
| The Global Health Observatory | https://www.who.int/data/gho/data/indicators |
| The Institute for Genome Sciences at the University of Maryland School of Medicine Genomic Center for Infectious Diseases (GCID) | https://gcid.igs.umaryland.edu/ |
| The Network Data Exchange (NDEx) | https://home.ndexbio.org/about-ndex/ |
| The Pan-Cancer Analysis of Whole Genomes (PCAWG) | NA |
| The World Reference Center for Emerging Viruses and Arboviruses | https://www.utmb.edu/wrceva |
| Therapeutically Applicable Research to Generate Effective Treatments initiative (TARGET) | https://www.cancer.gov/ccg/research/genome-sequencing/target#:~:text=The%20Therapeutically%20Applicable%20Research%20to,less%20toxic%20therapies%20for%20children. |
| Throughput Ranking by Iterative Analysis of Genomic Enrichment (TRIAGE) | https://www.niaid.nih.gov/research/triage |

| | |
|---|---|
| ToxoDB | https://toxodb.org/toxo/app |
| Trans-Omics for Precision Medicine (TOPMed) | https://topmed.nhlbi.nih.gov/ |
| Treehouse | https://treehousegenomics.soe.ucsc.edu/public-data/ |
| TrialShare | https://www.itntrialshare.org/ |
| TrichDB | https://trichdb.org/trichdb/app |
| TriTrypDB | https://tritrypdb.org/tritrypdb/app |
| Tuberculosis Regualory Network Analysis Tool (TBRNAT) | https://www.niaid.nih.gov/research/tuberculosis-regulatory-network-analysis-tool |
| UCSC Genome Browser | https://genome.ucsc.edu/ |
| UniProt | https://www.uniprot.org/ |
| United States Immunodeficiency Network (USIDNET) | https://usidnet.org/ |
| Vaccine Investigation and Online Information Network (VIOLIN) | https://violinet.org/introduction.php |
| VDJ Server | https://vdjserver.org/ |
| VDJbase | https://vdjbase.org/ |
| VectorBase | https://vectorbase.org/vectorbase/app |
| VEuPathDB | https://veupathdb.org/ |
| Virtual Biorepository Strain Catalog | https://arlgcatalogue.org/arlgCatalogue/ |
| Virus Pathogen Research (ViPR) | in BV-BRC |
| Vivli | https://vivli.org/ |
| Wake Forest Primate Studies Core | https://www.niaid.nih.gov/research/wake-forest-primate-studies-core |
| Women's Interagency HIV Study (WIHS) Public Dataset | https://www.niaid.nih.gov/research/womens-interagency-hiv-study |
| WorkflowHub | https://workflowhub.eu/ |
| WorldWide Antimalarial Resistance Network (WWARN) | https://www.iddo.org/wwarn |
| Worldwide Protein Databank | https://www.wwpdb.org/ |
| WormBase | https://wormbase.org//#012-34-5 |
| Yale Model Database | https://senselab.med.yale.edu/modeldb/ |
| Yale Protein Expression Database (YPED) | https://medicine.yale.edu/keck/nida/yped/ |
| Yoda Project | https://yoda.yale.edu/ |
| Zebrafish Information Network (ZFIN) | http://zfin.org/ |
| Zenodo | https://zenodo.org/ |
| AnVIL | https://anvilproject.org/data/consortia |
| BioData Catalyst | https://gen3.biodatacatalyst.nhlbi.nih.gov/ |
| NIH Helping to End Addiction Long-term (HEAL) Initiative Data Portal | https://healdata.org/portal/discovery |

| | |
|---|---|
| NCATS Biomedical Data Translator | https://ui.transltr.io/ |
| NCI Cancer Genomics Cloud | https://www.cancergenomicscloud.org/datasets |
| NCATS OpenData Portal | https://opendata.ncats.nih.gov/covid19 |
| Pathoplexus | https://pathoplexus.org/ |
| mapMECFS | https://mapmecfs.org/ |
